# A deep learning model for breast ductal carcinoma in situ classification in whole slide images

**DOI:** 10.1101/2022.01.14.22269329

**Authors:** Fahdi Kanavati, Shin Ichihara, Masayuki Tsuneki

## Abstract

The pathological differential diagnosis between breast ductal carcinoma in situ (DCIS) and invasive ductal carcinoma (IDC) is of pivotal importance for determining optimum cancer treatment(s) and clinical outcomes. Since conventional diagnosis by pathologists using microscopes is limited in terms of human resources, it is necessary to develop new techniques that can rapidly and accurately diagnose large numbers of histopathological specimens. Computational pathology tools which can assist pathologists in detecting and classifying DCIS and IDC from whole slide images (WSIs) would be of great benefit for routine pathological diagnosis. In this paper, we trained deep learning models capable of classifying biopsy and surgical histopathological WSIs into DCIS, IDC, and benign. We evaluated the models on two independent test sets (n=1,382, n=548), achieving ROC areas under the curves (AUCs) up to 0.960 and 0.977 for DCIS and IDC, respectively.

## 1 Introduction

According to the Global Cancer Statistics 2020Sung et al (2021), breast cancer is the fifth leading cause of cancer mortality worldwide with 685,000 deaths. Breast ductal carcinoma in situ (DCIS) encompasses heterogenous lesions defined as proliferation of neoplastic ductal epithelial cells within the mammary duct lobular systems. DCIS needs to be differentiated from benign lesions, such as usual ductal hyperplasia, and from precursor/risk lesions, like flat epithelial atypia or atypical ductal hyperplasia (ADH). In addition, there is a wide range of histopathological variety of DCIS from low-grade lesions to high-grade lesions which may have a risk for rapid progression to invasive ductal carcinoma (IDC) Collins et al (2004); Sanders et al (2015); Gupta et al (1997). The histopathological diagnosis of DCIS on core needle biopsy (CNB) specimen is generally accurate in both non-palpable mammographically detected lesions and palpable mass-forming tumors Dahlstrom et al (1996); Pijnappel et al (1997); El-Tamer et al (1999). Importantly, assessment of nuclear grading, architectural type, and the presence of necrosis in DCIS can be performed on CNB and is reasonably accurate and slightly similar to that of IDC Harris et al (2003). As for low-grade DCIS and smaller size CNB specimens, it might be associated with increased risk for a false-negative diagnosis in patients with DCIS Dillon et al (2006); on the other hand, false-positive diagnosis of DCIS in CNB is mainly caused by atypical ductal hyperplasia (ADH) Rakha and Ellis (2007). High-grade DCIS also has diagnostic problems; DCIS arising in sclerosing adenosis may be mistaken for IDC Eusebi et al (1989); Oberman and Markey (1991). Sometimes DCIS is found to arise near or in radial sclerosing lesions (so-called radial scar), and it becomes very difficult for pathologists to discriminate DCIS or IDC in tiny fragmented CNB specimens Bianchi et al (2012).

The definitive diagnosis of DCIS is extremely important because the presence or absence of associated invasion has a significant impact on prognosis Wapnir et al (2011); Thompson et al (2018). In addition, it has been recently suggested that some low-grade DCIS is indolent and not an immediate life-threatening condition Elshof et al (2018); Esserman et al (2014). The need for further stratification of heterogeneous DCIS has been pointed out, but interobserver agreement in grading is poor on Breast et al (1998); van Dooijeweert et al (2019). Conventional morphological diagnosis by human pathologists has limitations, and it is necessary to construct a new diagnostic strategy based on analysis of a large number of cases in the future. Genetic approaches using endogenous subtyping open up one possibility for new diagnostics, but it is also important to accumulate data discriminating DCIS and IDC using AI pathology diagnosis that provides highly reproducible morphological diagnostic data.

Deep learning has been widely applied in computational histopathology, with applications such as cancer classification in WSIs, cell detection and segmentation, and the stratification of patient outcomes Yu et al (2016); Hou et al (2016); Madabhushi and Lee (2016); Litjens et al (2016); Kraus et al (2016); Korbar et al (2017); Luo et al (2017); Coudray et al (2018); Wei et al (2019); Gertych et al (2019); Bejnordi et al (2017); Saltz et al (2018); Campanella et al (2019); Iizuka et al (2020). For breast histopathology in particular, deep learning has been applied for classification of cancer in WSIs Bayramoglu et al (2016); Sharma and Mehra (2020); Hameed et al (2020); Mi et al (2021); Sohail et al (2021); Wetstein et al (2021). As for histopathological grading of differentiation level for DCIS, a deep learning-based DCIS grading system that achieved a performance similar to expert observer pathologists was reported Wetstein et al (2021). IDC classification has also been recently investigated Kanavati and Tsuneki (2021a). Interestingly, magnification independent convolutional neural network (CNN) approach improved the performance of magnification specific model for breast cancer histopathology image classification Bayramoglu et al (2016).

In this study, we trained deep learning models (each consisting of a convolutional neural network (CNN) with/without an additional recurrent neural network (RNN)) to classify biopsy and surgical WSIs into DCIS, IDC, and benign. We evaluated the models on two different test sets obtained from different medical institutions achieving an ROC-AUC up to 0.960 and 0.977 for DCIS and IDC, respectively. These findings suggest that computational algorithms might be useful as routine histopathological diagnostic aids for in situ and invasive breast ductal carcinoma classification.

## 2 Materials and methods

### 2.1 Clinical cases and pathological records

This is a retrospective study. A total of 3,672 H&E (hematoxylin & eosin) stained histopathological specimens of human breast DCIS, IDC and benign lesions – 2,101 biopsy and 1,571 surgical – were collected from the surgical pathology files of two hospitals: International University of Health and Welfare, Mita Hospital (Tokyo) and Sapporo-Kosei General Hospital (Hokkaido) after histopathological review of those specimens by surgical pathologists. The cases were selected randomly so as to reflect a real clinical scenario as much as possible. The pathologists excluded cases that had poor scanned quality. Each WSI diagnosis was observed by at least two pathologists, with the final checking and verification performed by a senior pathologist. All WSIs were scanned at a magnification of x20 using the same Leica Aperio AT2 scanner and were saved SVS file format with JPEG2000 compression.

### 2.2 Dataset

Table 1 breaks down the distribution of the dataset into training, validation, and test sets. The split was carried out randomly taking into account the proportion of each label in the dataset. Hospitals which provided histopathological cases were anonymised (e.g., test set hospital 1 and hospital 2). The test sets were composed of WSIs of core needle biopsy and surgical specimens. The patients’ pathological records were used to extract the WSIs’ pathological diagnoses and to assign WSI labels. 100 WSIs out of the 299 WSIs with DCIS were manually annotated by pathologists to indicate loosely the cancerous tissue lesions. The rest of DCIS, IDC and benign WSIs were not annotated and the training algorithm only used the WSI diagnosis labels, meaning that the only information available for the training was whether the WSI contained DCIS, IDC, or benign, but no information about the location of the cancerous tissue lesions. If the WSI had both IDC and DCIS, the WSI label was considered to be IDC.

**Table 1:**
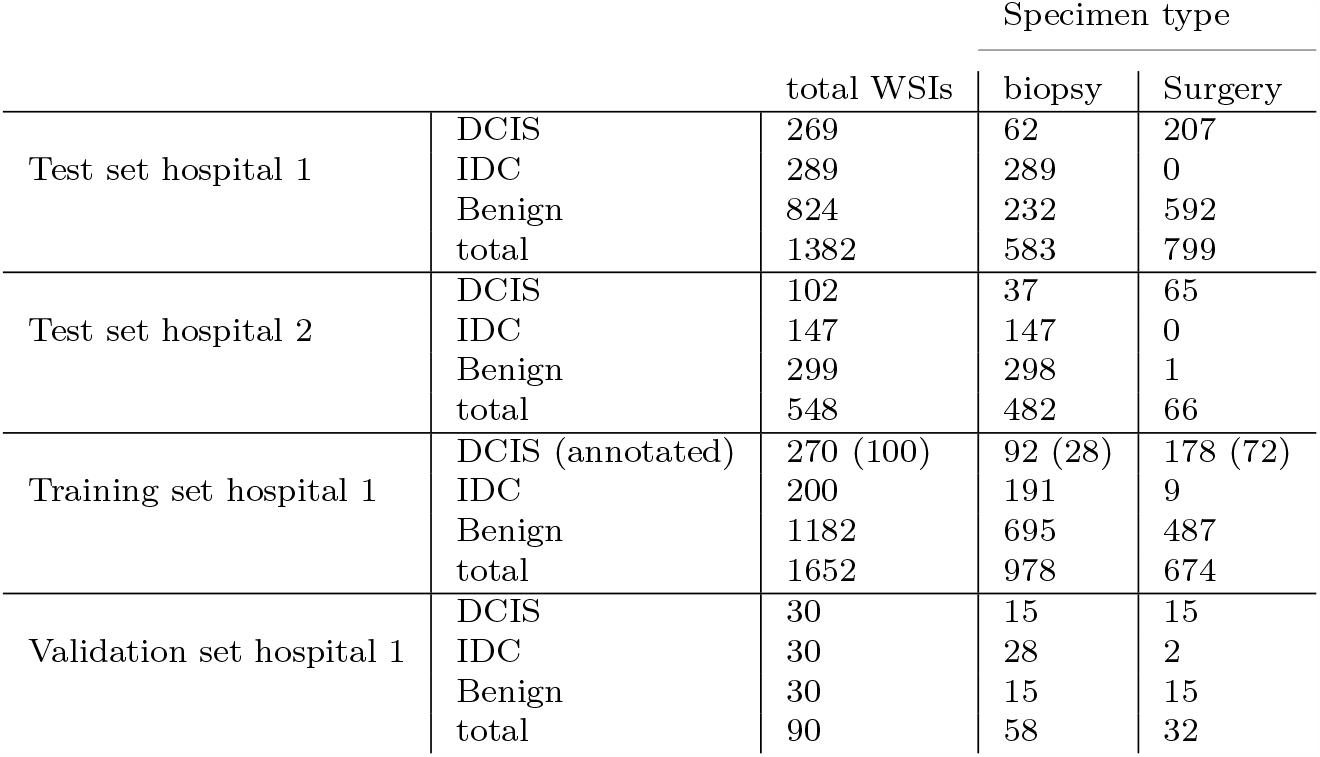
Distribution of the WSIs in the training, validation, and test sets.

### 2.3 Annotation

A senior pathologist, who performs routine histopathological diagnoses in general hospital, manually annotated 100 DCIS WSIs from the training set. The pathologist carried out coarse annotations by free-hand drawing (Fig. 3) using an in-house online tool developed by customising the open-source OpenSeadragon tool, which is a web-based viewer for zoomable images. On average, the pathologists annotated 5-6 lesions per WSI.

The pathologist performed annotations as follows: when scattered neoplastic ducts were observed, the pathologist annotated the myoepithelium at the limbus of the ducts (rather than only omitting the neoplastic epithelium) and the surrounding stromal collagen fiber that was compressed by the neoplastic duct (see Fig. 3 A). When DCIS was observed proliferating in clumps, the pathologist performed coarse annotations to take into account the irregularity of the margins, notch-like depressions, and the mixing of non-neoplastic ducts into the interior (black squares) (see Fig. 3 B). The pathologist included losses in the specimen due to calcification in the annotations except for coarse ones (see Fig. 3 C red arrowhead), and also included areas where fragmented epithelium was suspended within the necrotic material and if it was viable and neoplastic atypia could be identified (see Fig. 3 D). On the other hand, the pathologist omitted areas with large amount of necrosis in the ducts (see Fig. 3 C) and did not annotate areas where it was difficult to determine cytologically that the lesions were cancerous (see Fig. 3 E, F; blue arrowheads).

The average annotation time per WSI was about five minutes. Annotations performed by the pathologist were modified (if necessary), confirmed, and verified by two senior pathologists.

### 2.4 Deep learning models

Our deep learning models consisted of two separately-trained components: a CNN tile classifier and an RNN tile aggregator for WSI diagnosis. For the CNN, we have used the EfficientNetB1 architecture Tan and Le (2019) with a modified input size of 224×224px. We trained the models using the partial finetuning approach Kanavati and Tsuneki (2021b). This is where only the affine weights of the batch normalisation layers and the weights of final classification layer are updated while the rest remain frozen. We used as starting weights the pre-trained weights from ImageNet. The number of parameters that we fine-tuned in the model was 63,329.

In this study, we have used a similar training approach as that described in a previous study Kanavati et al (2020). The main difference, however, is that we used the partial fine-tuning method.

We started by detecting the tissue regions in the WSIs using Otsu’s method Otsu (1979) applied to the grayscale version of the WSIs. This allows the elimination of most of the white background. For training and inference, we needed then to extract tiles from the tissue regions. Instead of pre-extracting in advance, we extracted the tiles in real-time using the OpenSlide library Goode et al (2013). During inference, the slide tiling was done in a sliding window fashion on the tissue regions, using a fixed-size stride that was half the size of the tile. This means that when performing prediction on a WSI, we can obtain a regular grid-like output of predictions on the tissue regions, allowing us to visualise a heatmap of probability outputs which we can overlay on top of the WSI.

The training set consisted of a small set of annotated DCIS WSIs (n=100) and a large number of WSIs where only the WSI diagnosis was available as label. From the annotated WSIs, we only sampled tiles from the annotated tissue regions. Otherwise, we freely sampled tiles from the entire tissue regions.

During training, we maintained an equal balance of positive and negative labelled tiles in the training batch. To do so, we placed the WSIs in a shuffled queue with oversampling of the positive labels to ensure that all the WSIs were seen at least once during each epoch, and we looped over the labels in succession (i.e. we alternated between picking a WSI with a positive label – such as IDC and DCIS – and a negative label). Once a WSI was selected, we performed hard mining of tiles. To do so, the CNN was applied, in inference mode, in a sliding window fashion on all of the tissue regions if the WSI did not have annotations, or only on the annotated tissue regions if the WSI had annotations. We then selected the top *k* tiles with the highest probability for being positive. If the tile is from a negative WSI, this step effectively selects the false positives. The selected tiles were placed in a training subset, and once the number of tiles in the subset reached a given size *N*, a training pass was triggered. We used *k* = 4, *N* = 256, and a batch size of 32. Training thus alternated between inference and training sets.

The models were trained on WSIs at x5 and x10 magnifications. We used a tile size of 224×224px and a stride of 112×122px. During training, we performed real-time augmentation of the extracted tiles using variations of brightness, saturation and contrast. When using only the CNN, the WSI prediction was obtained by taking the maximum probability from all of the tiles.

We trained the CNN models with the Adam optimisation algorithm Kingma and Ba (2014) with the following parameters: *beta*_1_ = 0.9, *beta*_2_ = 0.999. We used a learning rate of 0.001. We applied a learning rate decay of 0.95 every 2 epochs. We used the binary cross entropy loss. We used early stopping by tracking the performance of the model on a validation set, and training was stopped automatically when there was no further improvement on the validation loss for 10 epochs. The model with the lowest validation loss was chosen as the final model.

An alternative approach to only taking the maximum probability of all tiles is the use of a RNN model. The RNN model can learn to combine the outputs from all the tiles to obtained a prediction for the WSI. We used an architecture with a single hidden layer of a gated recurrent unit (GRU) Cho et al (2014) with a size of 128 followed by a classification layer with a sigmoid activation and three outputs (DCIS, IDC, and benign). The inputs to the RNN model were the feature representation vectors obtained from the global-average pooling layer of the CNN classifier. All the feature vectors were fed into the RNN model to obtain the final WSI diagnosis. We extracted a set of feature vectors for all the WSIs in the training set using the CNN model as a feature extractor. Each WSI had a variable number of feature vectors and an associated WSI diagnosis. We trained the model with the Adam optimisation algorithm, with similar hyperparameters as the CNN training, except with a batch size of one.

### 2.5 Software and statistical analysis

The deep learning models were implemented and trained using Tensor-FlowAbadi et al (2015). AUCs were calculated in python using the scikit-learn packagePedregosa et al (2011) and plotted using matplotlib Hunter (2007). The 95% CIs of the AUCs were estimated using the bootstrap methodEfron and Tibshirani (1994) with 1000 iterations.

The true positive rate (TPR) was computed as

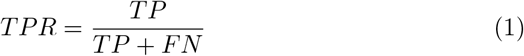

and the false positive rate (FPR) was computed as

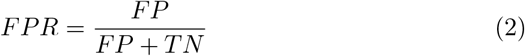

Where TP, FP, and TN represent true positive, false positive, and true negative, respectively. The ROC curve was computed by varying the probability threshold from 0.0 to 1.0 and computing both the TPR and FPR at the given threshold.

### 2.6 Availability of data and material

The datasets generated during and/or analysed during the current study are not publicly available due to specific institutional requirements governing privacy protection but are available from the corresponding author on reasonable request. The datasets that support the findings of this study are available from International University of Health and Welfare, Mita Hospital (Tokyo) and Sapporo-Kosei General Hospital (Hokkaido), but restrictions apply to the availability of these data, which were used under a data use agreement which was made according to the Ethical Guidelines for Medical and Health Research Involving Human Subjects as set by the Japanese Ministry of Health, Labour and Welfare, and so are not publicly available. However, the data are available from the authors upon reasonable request for private viewing and with permission from the corresponding medical institutions within the terms of the data use agreement and if compliant with the ethical and legal requirements as stipulated by the Japanese Ministry of Health, Labour and Welfare.

### 2.7 Code availability

To train the classification model in this study we used the publicly available TensorFlow training script available at https://github.com/tensorflow/models/tree/master/official/vision/image_classification.

## 3 Results

### 3.1 High AUC performance of WSI evaluation of breast DCIS and IDC histopathology images

The aim of this retrospective study was to train deep learning models for the classification of DCIS and IDC in WSIs. We trained a combination of CNN and RNN using a fast fine-tuning approach using a total of 1,652 WSIs for training. We evaluated the models on a two test sets each with 1,382 and 548 WSIs, respectively. For each test set, we computed the ROC AUC and log loss, and we have summarised the results in Tab. 2 and Fig. 2.

We trained using two different training approaches: (1) using the convolutional neural network (CNN) only and max-pooling the probabilities of all the tiles to obtain a WSI diagnosis; (2) using a CNN model to obtain the tile predictions followed by a recurrent neural network (RNN) to aggregate the tile predictions into a single WSI diagnosis (Fig. 1). We trained at two different magnifications x5 and x10. This resulted in three different models: WS x5; WS x10; WS x10 + RNN.

**Fig. 1:**
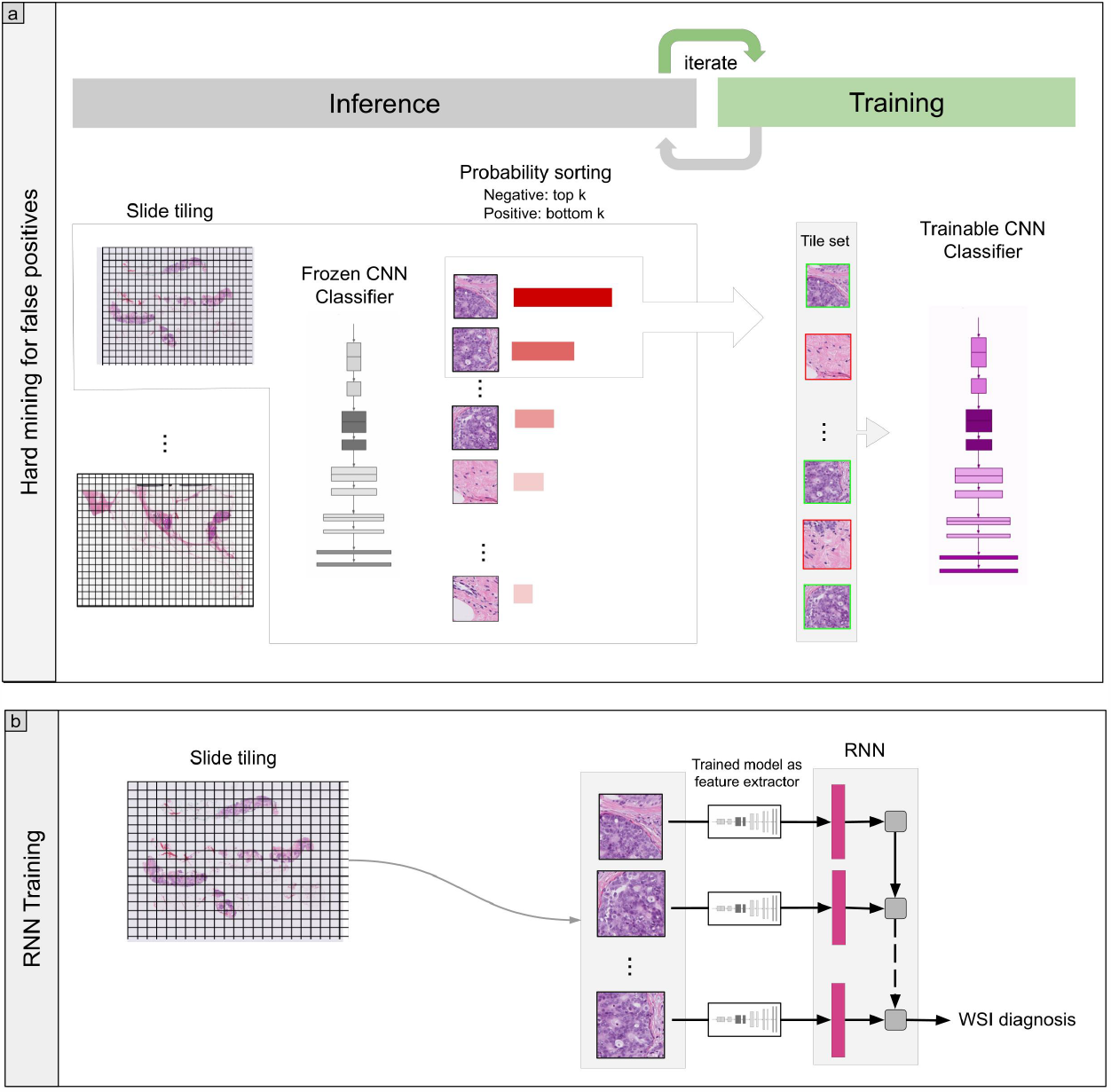
Overview of training method. (a) The training alternated between two steps: inference and training. During inference, the frozen model was applied in a sliding window fashion, and the top *k* tiles with highest probability were extracted. We maintained an equal balance of positive and negative labelled tiles in the training batch (see methods section for more details). The alter-nation between training and inference allows for hard mining of tiles. If the tile is from a negative WSI, this step effectively selects the false positives. The selected tiles were placed in a training subset, and once the number of tiles in the subset reached a given size *N*, the training step was triggered. (b) For the training of the RNN model, we used the trained CNN model to extract features from the WSIs in a sliding window fashion. These were then used for training the RNN model using only the WSI diagnosis label as output.

**Fig. 2:**
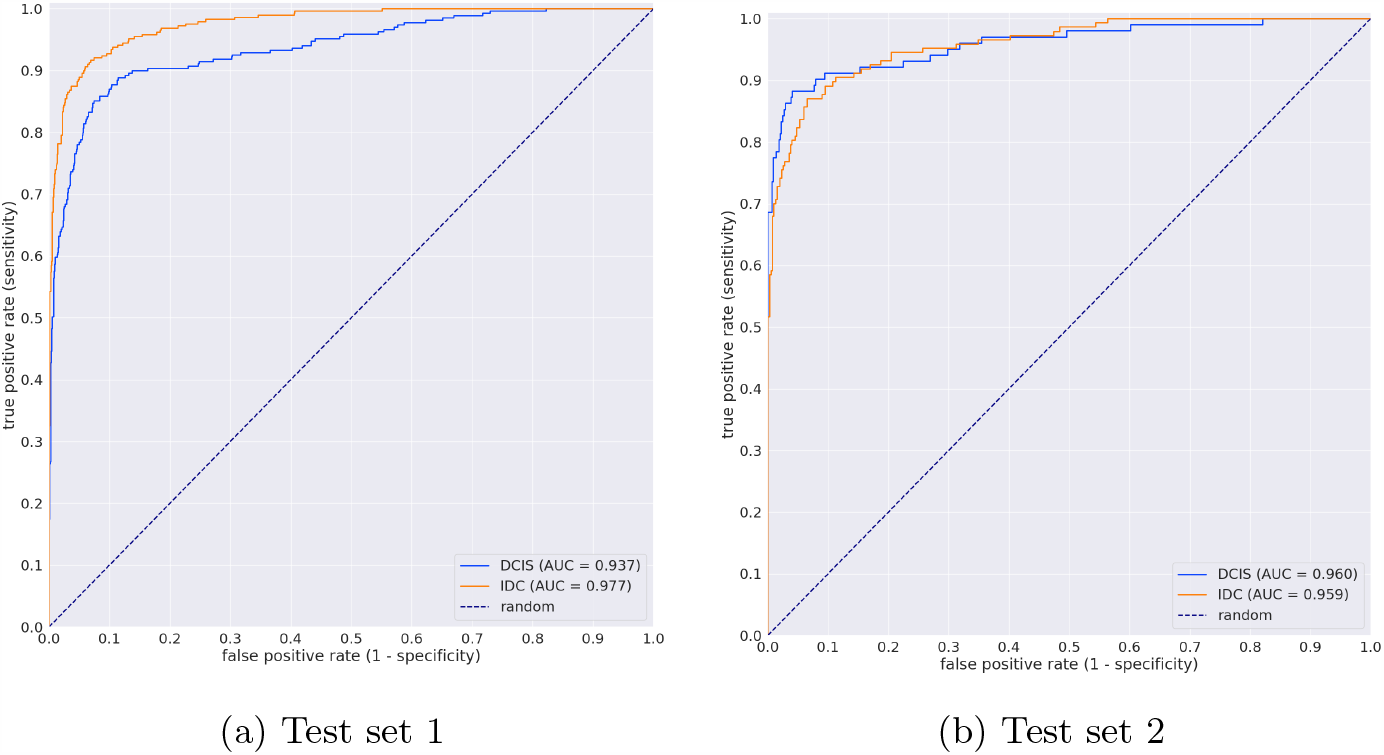
ROC curves with AUC for the two test sets

**Fig. 3:**
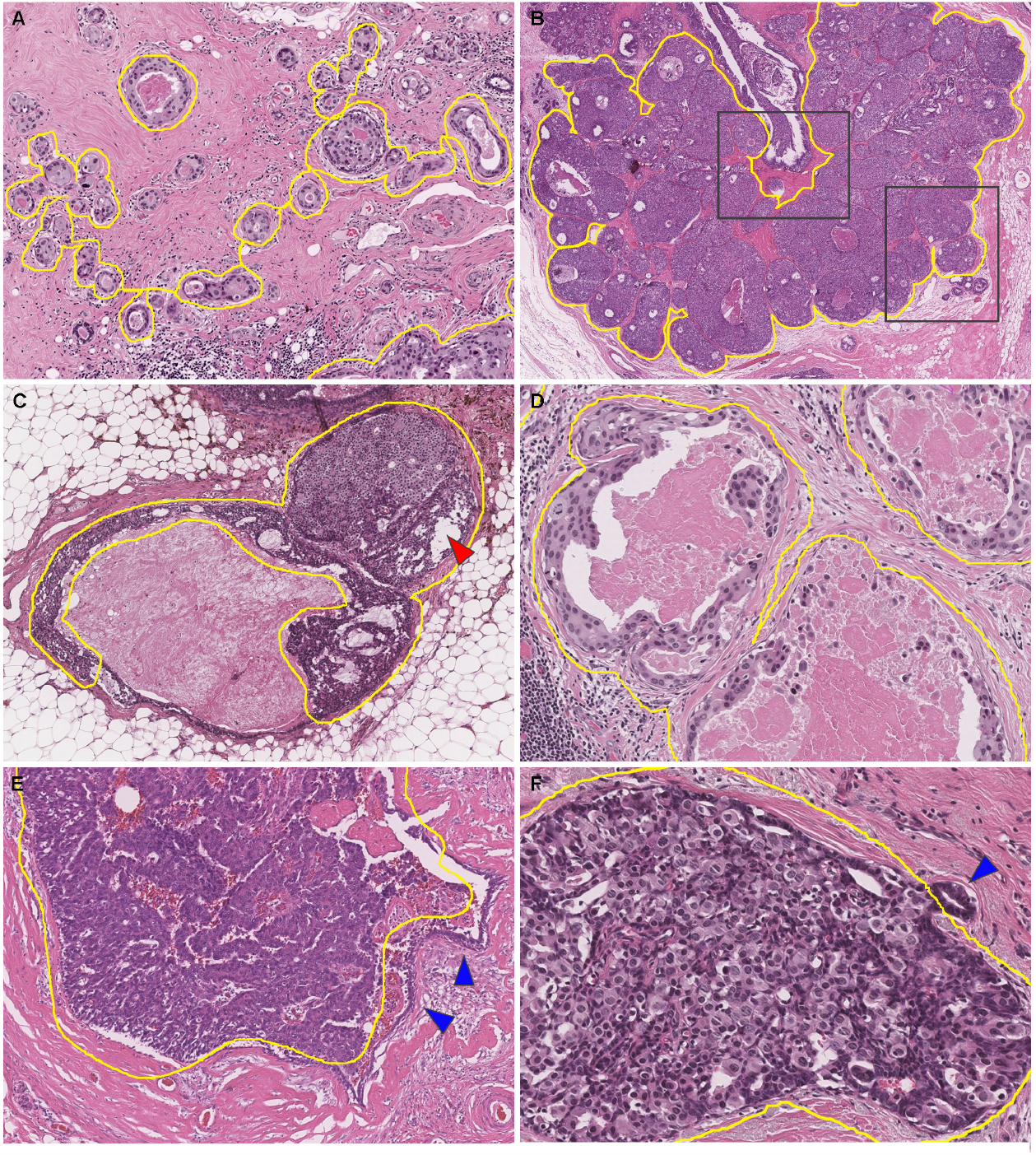
Representative manually drawing annotation images for breast ductal carcinoma in situ (DCIS). We performed annotations for each scattered neo-plastic ducts (A). We included the myoepithelium at the limbus of the ducts in the annotation area, rather than only omitting the neoplastic epithelium, and we also included the surrounding stromal collagen fiber that was compressed by the neoplastic duct. When annotating DCIS of the type that proliferates in clumps, we performed coarse annotations to take into account the irregularity of the margins, notch-like depressions, and the mixing of non-neoplastic ducts into the interior (black squares) (B). We omitted the areas with large amount of necrosis in the ducts (C). We included losses in the specimen due to calcification in the annotations except for coarse ones (red arrowhead) (C). We also included areas where fragmented epithelium was suspended within the necrotic material and if it was viable and neoplastic atypia could be identified (D). We did not annotate areas where it was difficult to determine cytologically that the lesions were cancerous (E, F; blue arrowheads).

We applied our models on two different test sets (Table 1). We have confirmed that surgical pathologists were able to diagnose these cases from visual inspection of the H&E stained slides alone. The model trained at x10 (WS x10) has a higher ROC-AUCs compared to the model trained at x5 (WS x5) for both DCIS and IDC (Table 2). The model (WS x10 + RNN) which consisted of CNN and RNN achieved highest ROC-AUCs of 0.960 (CI: 0.933 - 0.983) and 0.977 (CI: 0.967 - 0.984) for DCIS and IDC, respectively (Table 2). Figure 2 shows the ROC curves of on all test sets for each label (DCIS and IDC) from using the model (WS x10 + RNN).

**Table 2:**
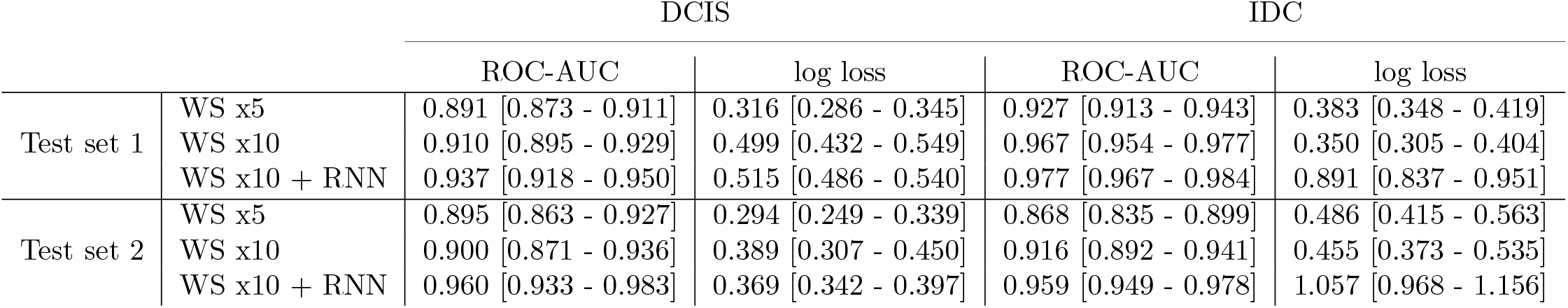
ROC AUC and log loss results for IDC and DCIS on the two test sets.

Figures 4, 5, and 6 show representative cases of true positive, false positive, and false negative, respectively from using the model (WS x10 + RNN).

**Fig. 4:**
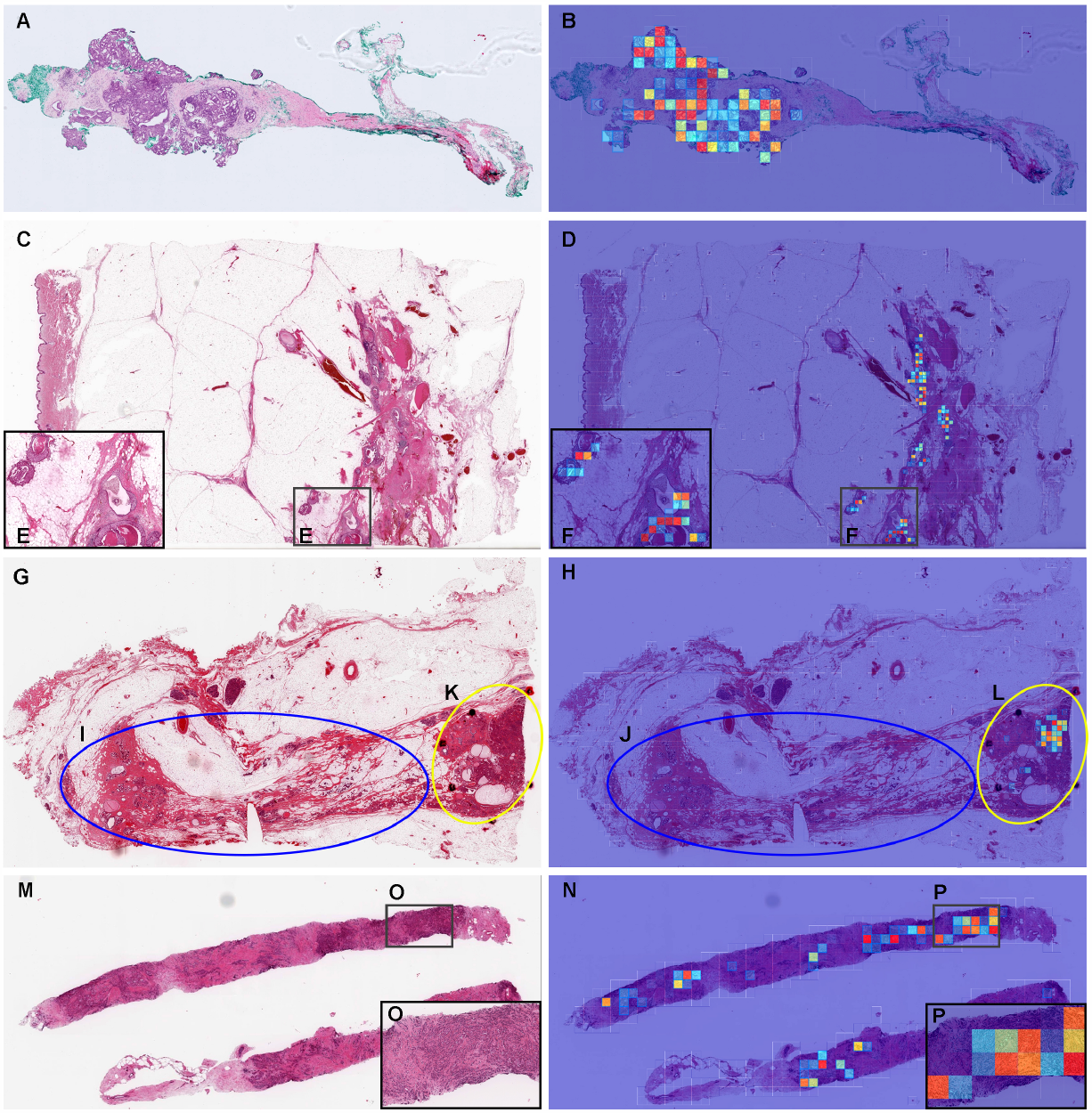
Representative true positive ductal carcinoma in situ (DCIS) and invasive ductal carcinoma (IDC) of breast from biopsy and surgical test sets. In the DCIS whole slide images (WSI) of biopsy (A) and surgical (C, G) specimens, the heatmap images show true positive prediction of DCIS cells (B, D, H) which correspond respectively to H&E histopathology (E, F, K, L). In many areas of surgical specimen (G), mastopathy/adenosis lesions are observed (I), and DCIS is observed at the margins (K) only. The heatmap image shows true negative (J) predictions of mastopathy/adenosis areas and true positive (L) predictions of DCIS areas. In the IDC WSI of biopsy (M), the heatmap image shows true positive predictions of IDC cells (N) which correspond respectively to H&E histopathology (O) and (P). The heatmap uses the jet color map where blue indicates low probability and red indicates high probability.

**Fig. 5:**
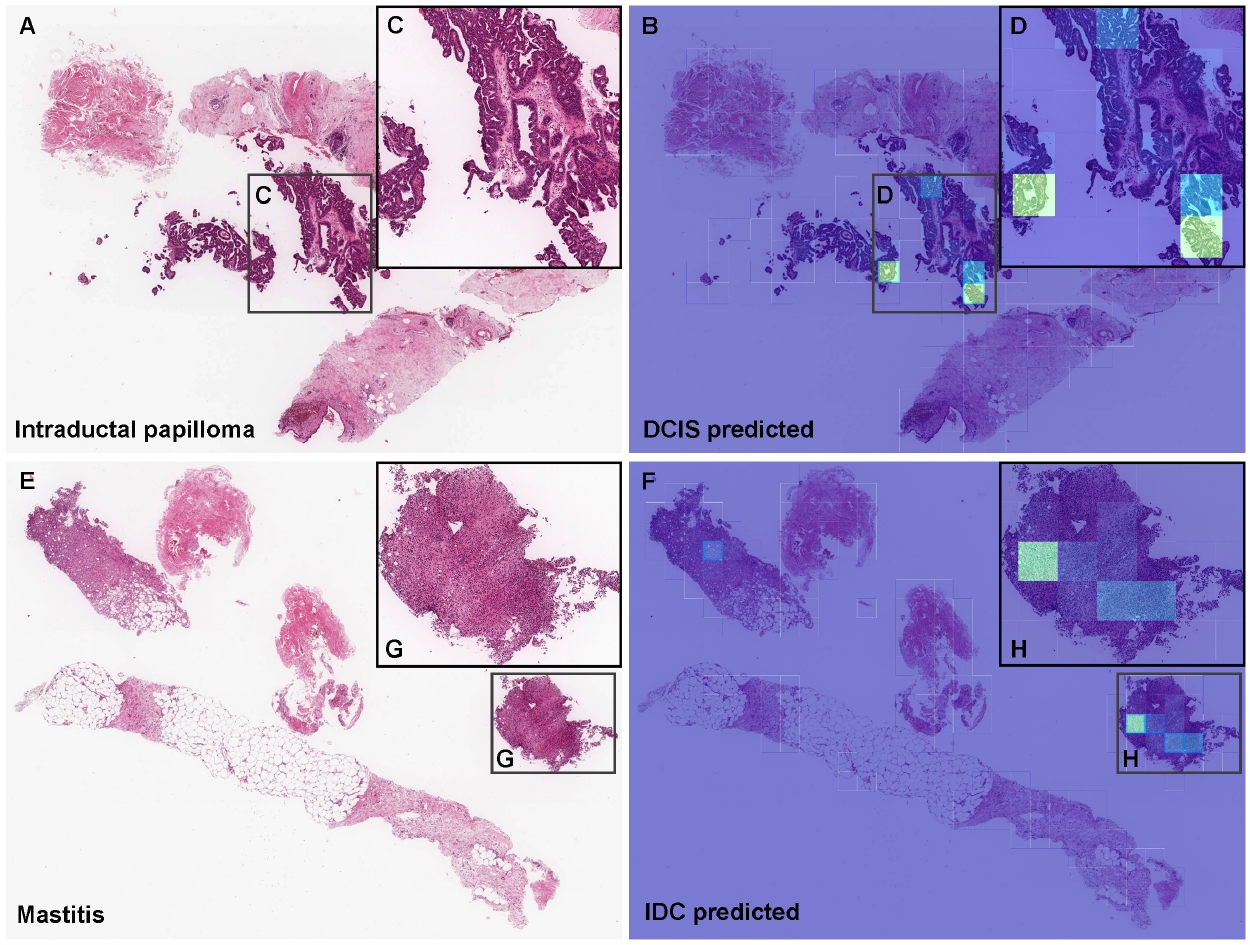
Representative examples of breast ductal carcinoma in situ (DCIS) and invasive ductal carcinoma (IDC) false positive prediction outputs on cases from biopsy test sets. Histopathologically, (A) and (E) are benign lesions (A: intraductal papilloma; E: Mastitis). The heatmap images (B, F) exhibited false positive predictions of DCIS (B) and IDC (F). Areas where specimen fragments and epithelium floated away from the stroma, with structural irregularity mimicking a cribriform patterns in the intraductal papilloma (C) would be the primary cause of false positive (D). Infiltration of chronic inflammatory cells (G) would be the primary cause of false positive due to its morphological analogous to IDC cells infiltration (H). The heatmap uses the jet color map where blue indicates low probability and red indicates high probability.

**Fig. 6:**
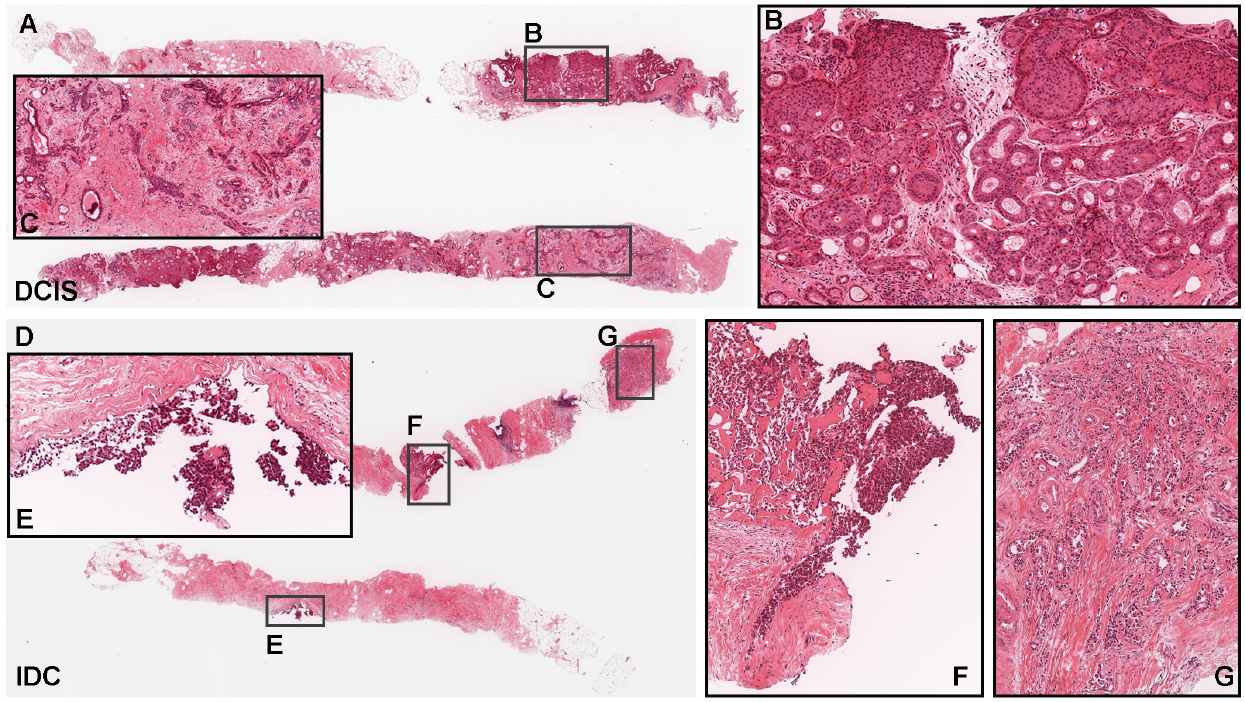
Representative false negative breast ductal carcinoma in situ (DCIS) and invasive ductal carcinoma (IDC) from biopsy test sets. According to the histopathological reports, this case (A) is a low-grade DCIS; however, oxiphilic changes in the cytoplasm and a low nuclear/cytoplasmic (N/C) ratio are demonstrated, reminiscent of DCIS with apocrine differentiation or neuroendocrine differentiation (B). In the background, mastopathic changes (C) are seen. Case (D) is intraductal carcinoma (E,F) with invasive component (G). Because the specimen is at the edge and has been destroyed, it is not easy for the pathologist to make a definite diagnosis, but small ducts with monotonous nuclei are infiltrating, suggesting IDC or tubular carcinoma.

### 3.2 True positive prediction

Our model satisfactorily predicted DCIS (Fig. 4B, D, F, H, L) and IDC (Fig. 4N, P) in biopsy (Fig. 4A, M) and surgical (Fig. 4D, H) specimens. The heatmap image shows true positive predictions (Fig. 4L) of DCIS areas (Fig. 4K) at surgical margin and true negative predictions (Fig. 4J) of mastopathy/adenosis areas which exhibit some similarity with DCIS and on occasion tends to be misdiagnosed (Fig. 4I).

### 3.3 False positive prediction

Histopathologically diagnosed intraductal papilloma (Fig. 5A, C) and mastitis (Fig. 5E, G) were false positively predicted for DCIS (Fig. 5B, D) and IDC (Fig. 5F, H), respectively.

### 3.4 False negative prediction

There were two types of false negatives: one was a low-grade DCIS with apocrine differentiation or neuroendocrine differentiation (Fig. 6A) and mastopathic changes in the background (Fig. 6C) based on eosinophilic changes in the cytoplasm and a low nuclear/cytoplasmic (N/C) ratio (Fig. 6B); the other (Fig. 6D) was DCIS (Fig. 6E, F) with invasive ductal carcinoma component (Fig. 6G) which was localized at the edge of the specimen. Both of these two cases were not easy for the pathologists to make a definitive diagnosis.

### 3.5 Controversial case

Interestingly, there was an IDC with focal invasive micropapillary carcinoma component predicted as DCIS (Fig. 7). In this case, the model using RNN had high probability predictions of both DCIS and IDC. It was reported that invasive micropapillary carcinoma growth pattern was present in 6% of all breast carcinomas Nassar et al (2001). While the diagnosis of invasive micropapillary carcinoma in most cases is obvious for pathologists, this particular cases presented a difficulty for the pathologists to diagnose based on H&E staining sections alone; this is because of the morphological similarities between IDC and DCIS in invasive micropapillary carcinoma in this particular case.

**Fig. 7:**
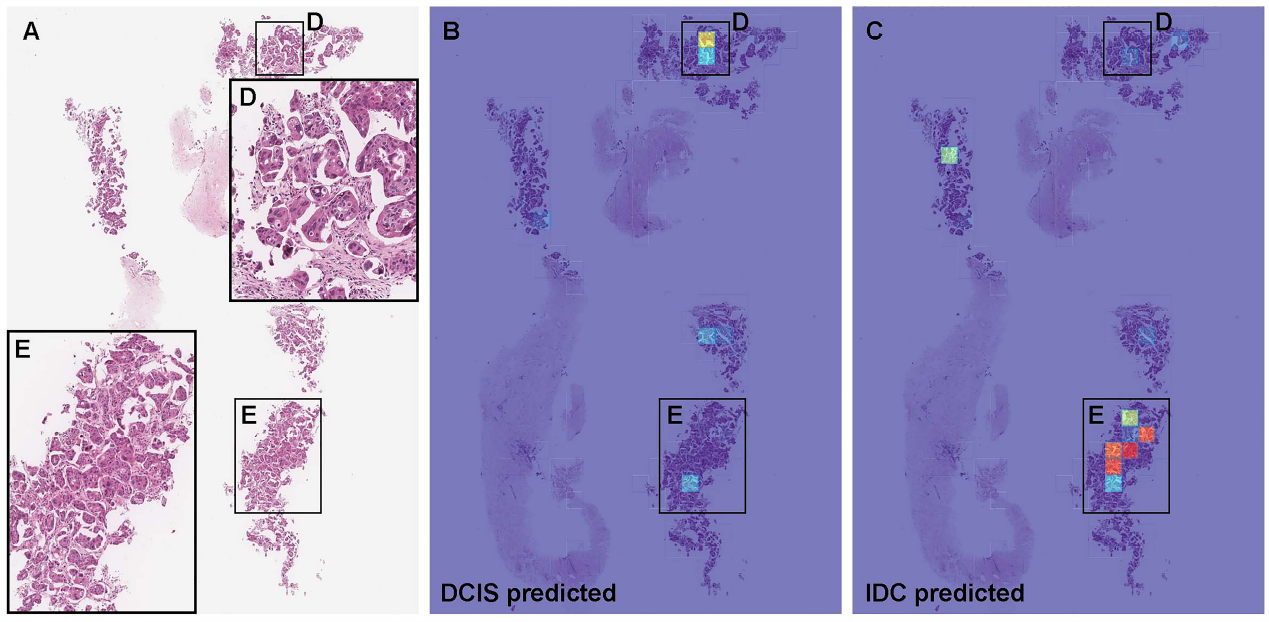
Representative controversial breast invasive ductal carcinoma (IDC) case. The true diagnosis of this case is IDC; however, it was predicted as breast ductal carcinoma in situ (DCIS). According to the histopathological report, this case is invasive micropapillary carcinoma (A). The heatmap images (B, C) exhibited false positive prediction of DCIS (B, D, E) and true positive prediction (C, D, E) in the same area. Based on the RNN model prediction, DCIS was predicted at whole slide image (WSI) level. The heatmap uses the jet color map where blue indicates low probability and red indicates high probability.

## 4 Discussion

In this study, we trained deep learning models for the classification of breast IDC and DCIS in surgical and biopsy WSIs. The deep learning models achieved AUCs in the range of 0.93-0.97.

When performing DCIS annotations, the pathologist avoided annotating the diverse non-neoplastic components that are often present in DCIS lesions. At the same time, the pathologist was careful to include the diverse range of varieties of DCIS features in the annotation areas (Fig. 3). In particular, the annotation areas included proliferating myoepithelial cells, which were surrounding ductal epithelial cells, and displaced stroma, which provides context information around the neoplastic tumor cells. The idea is that surgical pathologists consider not only the texture of the neoplastic epithelium (e.g., cell density, nuclear atypia, internuclear distance, cell homogeneity), but also the relationship between the neoplastic epithelial cell and its surroundings when performing diagnoses. As for discriminating between DCIS and microinvasive IDC, the presence or absence of microinvasion at the margins of the lesion is more important than cellular atypia within the lesion. It goes without saying that the key to assessing invasion is whether it extends beyond the myoepithelium and basement membrane; however, the inter-diagnostic concordance rate for the H&E stained section alone is not good, thus immunohistochemical staining is often helpful to reach definitive diagnosis Prasad et al (2000); Cserni et al (2016). In this study, it was revealed that the deep learning model could classify DCIS and IDC precisely by H&E section alone; it is conceivable that immunohistochemical staining might be omitted in the evaluation of the presence of ductal carcinoma cell invasion in the future.

Mastopathy and sclerosing lesions are often present in mammary tissue and it may become problematic to differentiate between benign and malignant Spruill (2016). It may be difficult to differentiate between sclerosing adenosis and IDC (stroma rich type), and radial sclerosing lesion (RSL) with epithelial atypia Rosa and Agosto-Arroyo (2019). In addition, apocrine metaplasia is also a common finding in both benign and malignant lesions; however, it is often difficult to evaluate nuclear atypia in lesions with apocrine metaplasia. During these differential diagnoses, surgical pathologists often perform immunohistochemistry to evaluate myoepithelium Hilson et al (2010); Tramm et al (2011) and grading of malignancies Moriya et al (2000); however, in rare cases accurate evaluation may become difficult using H&E staining alone or in combination with immunohistochemistry. In the present study, the deep learning model reached the high ROC-AUCs by using a wide variety of benign lesions (e.g., fibroadenoma, intraductal papilloma, adenosis, fibrocystic disease) as training sets for weakly supervision. Using such a wide variety of benign lesions would potentially be beneficial in training the model in expectation of potential cases it might encounter in a clinical setting and reduce the potential of false positive predictions. This could be important for other organs which have a wide variety of benign lesions, such as stomach (e.g., with a wide variety of gastric mucosal lesions, especially under H. pylori infection), liver (e.g., with a wide variety of changes due to viral hepatitis and NASH/NAFLD), and ovary (e.g., with many patterns of benign lesions).

A deep learning model (WS x10+RNN) established in the present study demonstrated high ROC-AUC performances to discriminate DCIS and IDC not only at WSI level but also tissue area level based on the obtained heatmaps. For example, in Fig. 4-G,H,I,J,K,L, the deep learning model predicted precisely not only the mastopathic changes in the background mammary glands as benign (non-neoplastic), but also specifically the DCIS invading areas in the mastopathy with dilated ducts (Fig. 4-K, L).

As for the false positives, most of the benign WSIs predicted as IDC had mastopathy in the tissue background. By reviewing heatmap images, especially, sparse growth of mesenchymal spindle cells and small ductal structures with inflammatory cell infiltration in the myxoidal stroma were tought to be the potential cause of false positives. In a core needle biopsy case with mastitis, it was predicted as IDC (Fig. 5-A, B). This might be due to the fact that core needle biopsy and/or surgery for mastitis cases were rarely performed in routine practice; therefore, there were not enough mastitis cases in the training sets. In the benign cases which were predicted as DCIS, there were isolated intraductal lesions at the edge of the CNB specimens (Fig. 5-A, B). In these cases, it was very difficult for surgical pathologists to differentiate DCIS from benign lesions (e.g., papilloma) based on only H&E staining section(s); therefore, the definitive diagnoses were made comprehensively based on the findings of immunohistochemistry Moriya et al (2009) and H&E histopathologies. It seems that benign lesions that mimic low-grade DCIS are difficult to classify by the deep learning model. However, in practical use, it is not weakness in AI-assisted diagnosis for lesions that are difficult to diagnose precisely for surgical pathologists.

There are two patterns of false negatives that we are interested in: one is DCIS with apocrine or neuroendocrine differentiation, and the other is stroma rich IDC with thin cord-like invasion Erber and Hartmann (2020) and tubular carcinoma with its extremely well differentiated structure (Fig. 6). The former shows eosinophilic cytoplasm, low N/C ratio, and irregular internuclear distance Coyne et al (2001); Damiani et al (1999). After histopathological reviewing, it was obvious that most of the false negative cases in the latter group exhibit morphology resembling sclerosing adenosis Becker et al (1992). It was reported that low grade DCIS represented the most common variant of breast in situ ductal carcinoma in sclerosing adenosis, and luminal A is the most predominant immunophenotype Yu et al (2018); Petersson et al (2010); Huang et al (2015). Thus, in the next step, it would be beneficial to perform iterative learning using enough number of DCIS WSIs with sclerosing adenosis. The deep learning models established in the present study achieved high ROC-AUC performances (Fig. 2 and Table 2); they offer promising results that indicate they could be beneficial as a screening aid for pathologists prior to observing histopathology on glass slides. In relatively large surgical specimens, it can be used as a double-check to reduce the risk of missed cancer foci. A deep learning model that can assist pathologists in classifying DCIS and IDC would be extremely beneficial for routine histopathological diagnosis. There was an issue with the model in differentiating between DCIS (low-grade) and benign lesions in this study. Since the prognostic significance and treatment methods might be different between low-grade and high-grade DCIS Elshof et al (2018); Esserman et al (2014), there might be an issue in including them in the same category as DCIS. However, at the same time, since there are large interobserver diagnostic differences in DCIS grading (low-grade and high-grade) on Breast et al (1998); van Dooijeweert et al (2019), there is an issue in collecting reliable training sets for supervised training. As future work, it would be worthwhile to establish deep learning models for predicting intrinsic Perou et al (2000); Sørlie et al (2001) or clinical Goldhirsch et al (2013); Coates et al (2015) subtypes based on H&E staining WSIs. Currently, surgical pathologists are able to predict clinical subtypes without immunohistochemistry to some extent based on the presence or absence of comedo-necrosis and the grading of nuclear atypia; however, deep learning models potentially can achieve similar predictive performance as surgical pathologists.

## Data Availability

Due to specific institutional requirements governing privacy protection, datasets used in this study are not publicly available.

## Acknowledgements

We are grateful for the support provided by Professor Takayuki Shiomi at Department of Pathology, Faculty of Medicine, International University of Health and Welfare; Dr. Ryosuke Matsuoka at Diagnostic Pathology Center, International University of Health and Welfare, Mita Hospital. We thank pathologists and oncologists who have been engaged in reviewing cases and clinicopathological discussion for this study.

## 6 Compliance with Ethical Standards

The experimental protocol was approved by the ethical board of the Sapporo-Kosei General Hospital (No. 580) and International University of Health and Welfare (No. 19-Im-007). All research activities complied with all relevant ethical regulations and were performed in accordance with relevant guidelines and regulations in the all hospitals mentioned above. Informed consent to use histopathological samples and pathological diagnostic reports for research purposes had previously been obtained from all patients prior to the surgical procedures at all hospitals, and the opportunity for refusal to participate in research had been guaranteed by an opt-out manner.

## 9 Contributions

F.K., S.I. and M.T. contributed equally to this study; F.K. and M.T. designed the studies; F.K., S.I., and M.T. performed experiments and analyzed the data; S.I. performed pathological diagnoses and reviewed cases; F.K. and M.T. performed computational studies; F.K., S.I., and M.T. wrote the manuscript; M.T. supervised the project. All authors reviewed and approved the final manuscript.

## References

Abadi M, Agarwal A, Barham P, et al (2015) TensorFlow: Large-scale machine learning on heterogeneous systems. URL https://www.tensorflow.org/, xsoftware available from tensorflow.org

Bayramoglu N, Kannala J, Heikkilä J (2016) Deep learning for magnification independent breast cancer histopathology image classification. In: 2016 23rd International conference on pattern recognition (ICPR), IEEE, pp 2440– 2445

Becker R, Mikel U, O’Leary T (1992) Morphometric distinction of sclerosing adenosis from tubular carcinoma of the breast. Pathology-Research and Practice 188(7):847–851

Bejnordi BE, Veta M, Van Diest PJ, et al (2017) Diagnostic assessment of deep learning algorithms for detection of lymph node metastases in women with breast cancer. Jama 318(22):2199–2210

Bianchi S, Giannotti E, Vanzi E, et al (2012) Radial scar without associated atypical epithelial proliferation on image-guided 14-gauge needle core biopsy: analysis of 49 cases from a single-centre and review of the literature. The Breast 21(2):159–164

on Breast ECWG, Sloane JP, Amendoeira I, et al (1998) Consistency achieved by 23 european pathologists in categorizing ductal carcinoma in situ of the breast using five classifications. Human pathology 29(10):1056–1062

Campanella G, Hanna MG, Geneslaw L, et al (2019) Clinical-grade computational pathology using weakly supervised deep learning on whole slide images. Nature medicine 25(8):1301–1309

Cho K, Van Merriënboer B, Gulcehre C, et al (2014) Learning phrase representations using rnn encoder-decoder for statistical machine translation. arXiv preprint 14061078

Coates AS, Winer EP, Goldhirsch A, et al (2015) Tailoring thera-pies—improving the management of early breast cancer: St gallen international expert consensus on the primary therapy of early breast cancer 2015. Annals of oncology 26(8):1533–1546

Collins L, Tamimi R, Baer H, et al (2004) Risk of invasive breast cancer in patients with ductal carcinoma in situ (dcis) treated by diagnostic biopsy alone: results from the nurses’ health study. Breast Cancer Research and Treatment 88

Coudray N, Ocampo PS, Sakellaropoulos T, et al (2018) Classification and mutation prediction from non–small cell lung cancer histopathology images using deep learning. Nature medicine 24(10):1559–1567

Coyne J, Dervan P, Barr L, et al (2001) Mixed apocrine/endocrine ductal carcinoma in situ of the breast coexistent with lobular carcinoma in situ. Journal of clinical pathology 54(1):70–73

Cserni G, Wells CA, Kaya H, et al (2016) Consistency in recognizing microinvasion in breast carcinomas is improved by immunohistochemistry for myoepithelial markers. Virchows Archiv 468(4):473–481

Dahlstrom J, Jain S, Sutton T, et al (1996) Diagnostic accuracy of stereotactic core biopsy in a mammographic breast cancer screening programme. Histopathology 28(5):421–427

Damiani S, Dina R, Eusebi V (1999) Eosinophilic and granular cell tumors of the breast. In: Seminars in diagnostic pathology, pp 117–125

Dillon M, Quinn C, McDermott E, et al (2006) Diagnostic accuracy of core biopsy for ductal carcinoma in situ and its implications for surgical practice. Journal of clinical pathology 59(7):740–743

van Dooijeweert C, van Diest PJ, Willems SM, et al (2019) Significant inter-and intra-laboratory variation in grading of ductal carcinoma in situ of the breast: a nationwide study of 4901 patients in the netherlands. Breast cancer research and treatment 174(2):479–488

Efron B, Tibshirani RJ (1994) An introduction to the bootstrap. CRC press

El-Tamer M, Axiotis C, Kim E, et al (1999) Accurate prediction of the amount of in situ tumor in palpable breast cancers by core needle biopsy: implications for neoadjuvant therapy

Elshof LE, Schmidt MK, Emiel JT, et al (2018) Cause-specific mortality in a population-based cohort of 9799 women treated for ductal carcinoma in situ. Annals of surgery 267(5):952

Erber R, Hartmann A (2020) Histology of luminal breast cancer. Breast Care 15(4):327–336

Esserman LJ, Thompson IM, Reid B, et al (2014) Addressing overdiagnosis and overtreatment in cancer: a prescription for change. The lancet oncology 15(6):e234–e242

Eusebi V, Collina G, Bussolati G (1989) Carcinoma in situ in sclerosing adenosis of the breast: an immunocytochemical study. In: Seminars in diagnostic pathology, pp 146–152

Gertych A, Swiderska-Chadaj Z, Ma Z, et al (2019) Convolutional neural networks can accurately distinguish four histologic growth patterns of lung adenocarcinoma in digital slides. Scientific reports 9(1):1483

Goldhirsch A, Winer EP, Coates A, et al (2013) Personalizing the treatment of women with early breast cancer: highlights of the st gallen international expert consensus on the primary therapy of early breast cancer 2013. Annals of oncology 24(9):2206–2223

Goode A, Gilbert B, Harkes J, et al (2013) Openslide: A vendor-neutral software foundation for digital pathology. Journal of pathology informatics 4

Gupta SK, Douglas-Jones AG, Fenn N, et al (1997) The clinical behavior of breast carcinoma is probably determined at the preinvasive stage (ductal carcinoma in situ). Cancer: Interdisciplinary International Journal of the American Cancer Society 80(9):1740–1745

Hameed Z, Zahia S, Garcia-Zapirain B, et al (2020) Breast cancer histopathology image classification using an ensemble of deep learning models. Sensors 20(16):4373. https://doi.org/10.3390/s20164373, URL https://doi.org/10.3390/s20164373

Harris GC, Denley HE, Pinder SE, et al (2003) Correlation of histologic prognostic factors in core biopsies and therapeutic excisions of invasive breast carcinoma. The American journal of surgical pathology 27(1):11–15

Hilson JB, Schnitt SJ, Collins LC (2010) Phenotypic alterations in myoepithelial cells associated with benign sclerosing lesions of the breast. The American journal of surgical pathology 34(6):896–900

Hou L, Samaras D, Kurc TM, et al (2016) Patch-based convolutional neural network for whole slide tissue image classification. In: Proceedings of the IEEE Conference on Computer Vision and Pattern Recognition, pp 2424– 2433

Huang N, Chen J, Xue J, et al (2015) Breast sclerosing adenosis and accompanying malignancies: a clinicopathological and imaging study in a chinese population. Medicine 94(49)

Hunter JD (2007) Matplotlib: A 2d graphics environment. Computing in Science & Engineering 9(3):90–95. https://doi.org/10.1109/MCSE.2007.55

Iizuka O, Kanavati F, Kato K, et al (2020) Deep learning models for histopathological classification of gastric and colonic epithelial tumours. Scientific reports 10(1):1–11

Kanavati F, Tsuneki M (2021a) Breast invasive ductal carcinoma classification on whole slide images with weakly-supervised and transfer learning. bioRxiv

Kanavati F, Tsuneki M (2021b) Partial transfusion: on the expressive influence of trainable batch norm parameters for transfer learning. arXiv preprint 210205543

Kanavati F, Toyokawa G, Momosaki S, et al (2020) Weakly-supervised learning for lung carcinoma classification using deep learning. Scientific reports 10(1):1–11

Kingma DP, Ba J (2014) Adam: A method for stochastic optimization. arXiv preprint 14126980

Korbar B, Olofson AM, Miraflor AP, et al (2017) Deep learning for classification of colorectal polyps on whole-slide images. Journal of pathology informatics 8

Kraus OZ, Ba JL, Frey BJ (2016) Classifying and segmenting microscopy images with deep multiple instance learning. Bioinformatics 32(12):i52–i59

Litjens G, Sánchez CI, Timofeeva N, et al (2016) Deep learning as a tool for increased accuracy and efficiency of histopathological diagnosis. Scientific reports 6:26, 286

Luo X, Zang X, Yang L, et al (2017) Comprehensive computational pathological image analysis predicts lung cancer prognosis. Journal of Thoracic Oncology 12(3):501–509

Madabhushi A, Lee G (2016) Image analysis and machine learning in digital pathology: Challenges and opportunities. Medical Image Analysis 33:170– 175

Mi W, Li J, Guo Y, et al (2021) Deep learning-based multi-class classification of breast digital pathology images. Cancer Management and Research Volume 13:4605–4617. https://doi.org/10.2147/cmar.s312608, URL https://doi.org/10.2147/cmar.s312608

Moriya T, Sakamoto K, Sasano H, et al (2000) Immunohistochemical analysis of ki-67, p53, p21, and p27 in benign and malignant apocrine lesions of the breast: its correlation to histologic findings in 43 cases. Modern Pathology 13(1):13–18

Moriya T, Kozuka Y, Kanomata N, et al (2009) The role of immunohistochemistry in the differential diagnosis of breast lesions. Pathology 41(1):68–76

Nassar H, Wallis T, Andea A, et al (2001) Clinicopathologic analysis of invasive micropapillary differentiation in breast carcinoma. Modern Pathology 14(9):836–841

Oberman H, Markey B (1991) Noninvasive carcinoma of the breast presenting in adenosis. Modern Pathology 4(1):31–35

Otsu N (1979) A threshold selection method from gray-level histograms. IEEE transactions on systems, man, and cybernetics 9(1):62–66

Pedregosa F, Varoquaux G, Gramfort A, et al (2011) Scikitlearn: Machine learning in Python. Journal of Machine Learning Research 12:2825–2830

Perou CM, Sørlie T, Eisen MB, et al (2000) Molecular portraits of human breast tumours. nature 406(6797):747–752

Petersson F, Tan PH, Choudary Putti T (2010) Low-grade ductal carcinoma in situ and invasive mammary carcinoma with columnar cell morphology arising in a complex fibroadenoma in continuity with columnar cell change and flat epithelial atypia. International journal of surgical pathology 18(5):352–357

Pijnappel RM, van Dalen A, Rinkes IHB, et al (1997) The diagnostic accuracy of core biopsy in palpable and non-palpable breast lesions. European journal of radiology 24(2):120–123

Prasad ML, Osborne MP, Giri DD, et al (2000) Microinvasive carcinoma (t1mic) of the breast: clinicopathologic profile of 21 cases. The American journal of surgical pathology 24(3):422–428

Rakha E, Ellis I (2007) An overview of assessment of prognostic and predictive factors in breast cancer needle core biopsy specimens. Journal of clinical pathology 60(12):1300–1306

Rosa M, Agosto-Arroyo E (2019) Core needle biopsy of benign, borderline and in-situ problematic lesions of the breast: Diagnosis, differential diagnosis and immunohistochemistry. Annals of diagnostic pathology 43:151, 407

Saltz J, Gupta R, Hou L, et al (2018) Spatial organization and molecular correlation of tumor-infiltrating lymphocytes using deep learning on pathology images. Cell reports 23(1):181–193

Sanders ME, Schuyler PA, Simpson JF, et al (2015) Continued observation of the natural history of low-grade ductal carcinoma in situ reaffirms proclivity for local recurrence even after more than 30 years of follow-up. Modern pathology 28(5):662–669

Sharma S, Mehra R (2020) Conventional machine learning and deep learning approach for multi-classification of breast cancer histopathology images—a comparative insight. Journal of Digital Imaging 33(3):632–654. https://doi.org/10.1007/s10278-019-00307-y, URL https://doi.org/10.1007/s10278-019-00307-y

Sohail A, Khan A, Nisar H, et al (2021) Mitotic nuclei analysis in breast cancer histopathology images using deep ensemble classifier. Medical Image Analysis 72:102, 121. https://doi.org/10.1016/j.media.2021.102121, URL https://doi.org/10.1016/j.media.2021.102121

Sørlie T, Perou CM, Tibshirani R, et al (2001) Gene expression patterns of breast carcinomas distinguish tumor subclasses with clinical implications. Proceedings of the National Academy of Sciences 98(19):10,869–10,874

Spruill L (2016) Benign mimickers of malignant breast lesions. In: Seminars in diagnostic pathology, Elsevier, pp 2–12

Sung H, Ferlay J, Siegel RL, et al (2021) Global cancer statistics 2020: Globocan estimates of incidence and mortality worldwide for 36 cancers in 185 countries. CA: a cancer journal for clinicians 71(3):209–249

Tan M, Le Q (2019) Efficientnet: Rethinking model scaling for convolutional neural networks. In: International Conference on Machine Learning, PMLR, pp 6105–6114

Thompson AM, Clements K, Cheung S, et al (2018) Management and 5-year outcomes in 9938 women with screen-detected ductal carcinoma in situ: the uk sloane project. European Journal of Cancer 101:210–219

Tramm T, Kim JY, Tavassoli FA (2011) Diminished number or complete loss of myoepithelial cells associated with metaplastic and neoplastic apocrine lesions of the breast. The American journal of surgical pathology 35(2):202– 211

Wapnir IL, Dignam JJ, Fisher B, et al (2011) Long-term outcomes of invasive ipsilateral breast tumor recurrences after lumpectomy in nsabp b-17 and b-24 randomized clinical trials for dcis. Journal of the National Cancer Institute 103(6):478–488

Wei JW, Tafe LJ, Linnik YA, et al (2019) Pathologist-level classification of histologic patterns on resected lung adenocarcinoma slides with deep neural networks. Scientific reports 9(1):1–8

Wetstein SC, Stathonikos N, Pluim JPW, et al (2021) Deep learning-based grading of ductal carcinoma in situ in breast histopathology images. Laboratory Investigation 101(4):525–533. https://doi.org/10.1038/s41374-021-00540-6, URL https://doi.org/10.1038/s41374-021-00540-6

Yu BH, Tang SX, Xu XL, et al (2018) Breast carcinoma in sclerosing adenosis: a clinicopathological and immunophenotypical analysis on 206 lesions. Journal of clinical pathology 71(6):546–553

Yu KH, Zhang C, Berry GJ, et al (2016) Predicting non-small cell lung cancer prognosis by fully automated microscopic pathology image features. Nature communications 7:12, 474

